# Knowledge and Perception of COVID-19, Prevalence of Pre-Existing Conditions, and Access to Essential Resources and Health Services in Somali IDP Camps

**DOI:** 10.1101/2020.08.17.20176271

**Authors:** Jude Alawa, Samir Al-Ali, Lucas Walz, Eleanor Wiles, Nikhil Harle, Mohamed Abdullahi Awale, Deqo Mohamed, Kaveh Khoshnood

**Author notes:** These authors contributed equally to this work (Samir Al-Ali, Lucas Walz). Authors listed alphabetically. Correspondence to: Dr. Kaveh Khoshnood, 60 College Street, Ste 826, New Haven, CT, 06510, USA.

## Abstract

**Background:** Three million internally displaced Somalis live in overcrowded settlements with weakened infrastructure, insufficient access to WASH facilities, and inaccessible health services. This vulnerable population is especially susceptible to COVID-19, which is expected to have worsened health outcomes and exacerbate existing structural challenges in the implementation of public health measures. This study examines knowledge of COVID-19, self-reported prevalence of preexisting conditions, and access to essential health services among residents of internally displaced persons (IDP) camps in Somalia.

**Methods:** A descriptive, cross-sectional survey design assessing demographics, current health profiles, knowledge and perceptions of COVID-19, and access to resources was used. 401 Somali IDP camp residents completed the survey.

**Results:** Though 77% of respondents reported taking at least one COVID-19 preventative public health measure, respondents reported a severe lack of access to adequate sanitation, an inability to practice social distancing, and nearly universal inability to receive a COVID-19 screening exam. Questions assessing knowledge surrounding COVID-19 prevention and treatment yielded answers of “I don’t know” for roughly 50% of responses. The majority were not familiar with basic information about the virus or confident that they could receive medical services if infected. Those who perceived their health status to be “fair,” as opposed to “good,” showed 5.69 times higher odds of being concerned about contracting COVID-19. Respondents who felt more anxious or nervous and those who introduced one behavioral change to protect against COVID-19 transmission showed 10.16 and 5.20 times increased odds of being concerned about disease contraction, respectively.

**Conclusion:** This study highlights immense gaps in the knowledge and perceptions of COVID-19 and access to treatment and preventative services among individuals living in Somali IDP camps. A massive influx of additional resources is required to adequately address COVID-19 in Somalia, starting with educating those individuals most vulnerable to infection.

**What is already known?:**

- There are no studies to date examining COVID-19 symptoms, as well as attitudes and perceptions, in Somali IDP camps.
- Investigations performed in other camp-like humanitarian settings have demonstrated high prevalence of COVID-19 symptoms.

**What are the new findings?:**

- People living in Somali IDP camps are generally unfamiliar with basic COVID-19 information, such as the possibility of transmission by asymptomatic individuals.
- A majority of respondents displayed at least one symptom consistent with COVID-19, and the vast majority were unable to access COVID-19 screening services.
- Older age, having implemented one behavioral change to protect against contraction, new stress or anxiety, and a “fair” health perception were significant predictors of being concerned about contracting COVID-19.

**What do the new findings imply?:**

- There is a significant gap in the knowledge and perception of COVID-19 by those in Somali IDP camps.
- Utilizing trusted sources of information may be an effective way of disseminating COVID-19 related information among Somali IDP camp residents.

## Introduction

There are more than 41 million internally displaced persons (IDPs) and 25 million refugees around the world according to the United Nations High Commissioner for Refugees (UNHCR)[1]. Many of these displaced people live in large-scale camps with conditions that could undermine public health guidelines for the control of COVID-19, such as social distancing, and are thus especially vulnerable to emerging infectious diseases[2]. Additional legal, financial, and linguistic barriers often inhibit displaced residents from accessing their host country’s health care system, exacerbating conditions precipitated by an absence of basic amenities, such as soap, running water, and medical personnel[2]. Eighty-four percent of displaced people reside in low- or middle-income countries, and these host countries are often ill-equipped to rapidly institute health management protocols and response teams within IDP communities, especially amidst the COVID-19 pandemic[3]. Providing continuity of care for chronic conditions as well as infection prevention and control measures within displaced communities are necessary to ensure the well-being of both displaced and host communities[2, 4].

The Ministry of Health in Somalia announced the first confirmed case of COVID-19 in Somalia on March 16, 2020[5]. As of July 8, 2020, the Ministry of Health of Somalia reported 3,015 confirmed COVID-19 cases, 1,827 of which are active and 92 of which have resulted in death[5]. Although the prevalence of COVID-19 remains low at the moment, the nation’s health system is extremely underprepared for a further outbreak of COVID-19[6].

In Somalia, thirty years of civil war and natural disasters have resulted in nearly three million internally displaced persons who now inhabit over 2,100 overcrowded settlements[7]. The prolonged conflict has severely damaged Somalia’s healthcare infrastructure, leaving its inhabitants vulnerable to climate related disasters, malnourishment, infectious disease outbreaks, and other humanitarian crises[8, 9].

Somalis face several barriers in utilizing their country’s healthcare system. Several reports inculcate that health worker shortages and a nonexistent health information system leave its residents without reliable access to care[8, 9]. Somali physicians have also reported that there are no ventilators and only two intensive care units with a total of 31 beds across the country, which is alarming given that these resources are regarded as necessary to treat severe COVID-19 infections[10]. In addition, it has been reported that two in three Somalis have difficulties accessing safe water and approximately 50% of health care centers do not have reliable access to electricity[11, 12]. Qualitative reports also signal that care-seeking behavior is very poor among Somalis, as a result of distrust towards the system’s lack of regulation and unaffordable prices[13]. Consequently, Somalia has some of the lowest health indicators in the world with life expectancies of 54 years for males and 57 years for females[11, 14]. Studies in Somalia have reinforced a predisposition of internally displaced persons to infectious and water-borne diseases[15]. IDPs are among the most vulnerable to infectious disease outbreaks as they face circumstances such as overcrowding, uncontained sewage and waste, limited access to health services, contaminated water, low immunization coverage, and stigmatization.

According to the Global Health Security Index, Somalia ranks 194 out of 195 countries in preparedness for a globally catastrophic biological event[16]. To address the dire threat COVID- 19 poses to IDPs in Somalia, ongoing efforts have primarily focused on scaling up water, sanitation, and hygiene (WASH) services, increasing monitoring, and disseminating information about COVID-19[6, 7, 17–19]. Many organizations have also helped obtain equipment such as personal protective equipment (PPE) and ventilators for providers, as well as soap and chlorine tablets for community members[7, 10, 19]. Several organizations have reported that the country’s health system has no capacity to make early case detections, isolate and care for patients, and trace contacts, and have thus focused their efforts on preventive measures to mitigate the virus’s impact[13].

Among these efforts, few have incorporated IDP knowledge and perceptions of COVID-19 in their planning and implementation. However, studies on Bangladeshi IDP camps similar to those in Somalia warn of the profound consequences of COVID-19 on those without a strong healthcare infrastructure[20]. The scarcity of IDP-focused COVID-19 data has not prevented various international health authorities from warning of the grave threats the pandemic poses to these vulnerable populations[21, 22]. Nevertheless, there is an international information gap about IDP knowledge and perceptions that inhibit providers from implementing sustainable, culturally appropriate health interventions.

This study aims to explore knowledge and perceptions of the symptoms, transmission, prevention, and treatment of COVID-19 among persons living in IDP camps and to understand the unique structural barriers that inhibit a comprehensive public health response in this setting. We hope that our study findings contribute to the development of potential interventions to improve the response to COVID-19 in Somali IDP camps, where some of the world’s most vulnerable individuals reside.

## Methods

### Design and Instrument

A descriptive, cross-sectional survey tool was used to assess the current health profile, living conditions, and knowledge and perceptions surrounding COVID-19 among adults living in Somali IDP camps. To design the survey tool, guidance was drawn primarily from published WHO and CDC information on COVID-19 symptoms and transmission, a 2019 Displacement Severity Assessment among forcibly evicted Somalis conducted by the Internal Displacement Monitoring Centre, and two previously published survey studies which evaluated COVID-19 knowledge and risk factors in limited-resource settings[23–27].

Our adapted survey tool begins with a section collecting demographic information on participants, including sex, age, displacement status, geographic location of their camp, household size and status, education, and employment. Responses for educational status reflect the education system in Somalia, and included options for “no formal education” and for all levels from primary through tertiary schools, as well as options for graduate degrees and Qur’anic education. The next section of the survey gathers information on participants’ health profile, with questions on self-rating of current health, existing conditions, and current symptoms if applicable. This section also employs a 12-item tool to gauge concerns brought on by COVID- 19, such as potential effects on mental wellbeing, contracting COVID-19, or ability to buy essential food items. The third section of the survey measures knowledge of COVID-19, most commonly used sources of information, and trust in various knowledge sources and community efforts against COVID-19. An open-ended question on COVID-19 knowledge was followed by a 23-item tool which assesses knowledge of COVID-19, and consists of true-or-false statements regarding key facts or misconceptions surrounding the disease. The final section of the survey evaluates access to medical resources, COVID-19 tests and treatments, and essential services.

Because many participants did not speak English, this survey tool was translated into Somali. The Somali version of the survey was then reviewed for its content and suitability in the given context. Minor changes to the language of the Somali version were made to improve readability.

### Sample and Setting

A convenience sample of 401 individuals living in twelve Somali IDP camps across six areas (Ceelasha, Lafoole, Xaawo Cabdi, Carbiska, and Afgooye) of the Lower Shabelle region in Somalia, often referred to as the world’s capital of IDPs, was obtained in June 2020. Participants must have been older than 18 years of age, physically able to complete the survey, and willing to take part in the study. If they agreed to be included in the study, a brief presentation of the purpose, procedure, and requirements for participation was given privately. Prior to the survey being interviewer-administered by multilingual and trained staff from the Hagarla Institute, verbal consent was obtained from each participant. The Hagarla Institute is a non-profit organization dedicated to furthering clinical research, capacity-building, and skills transfer for medical personnel across Africa. Participants were informed that they had the right to withdraw at any time and that there would be no consequences for withdrawal. All information collected was kept confidential and anonymized by removing all identifiable information. This study received approval from the ethics board at SIMAD University in Somalia and was deemed exempt from review by the Yale IRB (ID #2000028344).

### Data Analysis

Responses from each survey were manually input onto Qualtrics survey software in English and analyzed using SAS Studio 3.8[28]. Sample descriptive statistics were used to report median and standard deviation calculations for continuous variables, along with frequency and percentages of responses for categorical variables. Bivariate and multivariate logistic regressions were also conducted to demonstrate the relationships between concern about contracting COVID-19 and sex, age (dichotomized), education, food access, employment status, pre-existing conditions, symptoms present, actions taken, and healthcare utilization. These findings are presented with 95% confidence intervals.

## Results

### Sample Demographics

The demographic characteristics of survey participants are shown in **Table 1**. Of the 401 eligible participants who recorded responses, 382 (96%) identified as IDPs and 15 (4%) identified as refugees. The survey had a completion rate of 99%. The vast majority of the participants were female (86%, n = 344) and the median age of the participants was 32.0, with a standard deviation of 13.2 years. Most participants either had no formal education (89%, n = 353) or received Qur’anic education (32%, n = 126), with only 10 participants (3%) having received education past secondary school.

**Table 1.** Respondent Characteristics.

| Characteristic | N (%)<sup>a,b</sup> |
| --- | --- |
| **Sex** |  |
| Female | 344 (86.2) |
| Male | 55 (13.8) |
| **Age** | 32.0 ± 13.2 |
| **Status** |  |
| IDP | 382 (96.2) |
| Refugee | 15 (3.8) |
| **Area** |  |
| Bismillah | 46 (11.7) |
| Iskaashi | 146 (37.6) |
| Marko | 37 (10.0) |
| Other | 167 (40.7) |
| **# of People in Household** | 7 ± 5.1 |
| **# of People in Household over 65** | 0 ± 3.0 |
| **House or Dwelling** |  |
| Own | 97 (24.3) |
| Rent | 38 (9.5) |
| Not Sure | 265 (66.3) |
| **Highest Level of Education** |  |
| No Formal Education | 353 (88.7) |
| Primary School | 26 (6.5) |
| Lower Secondary School | 5 (1.3) |
| Upper Secondary School | 4 (1.0) |
| Post-secondary/tertiary school | 7 (1.8) |
| Graduate Degree (e.g. Master's, Doctorate) | 3 (0.8) |
| Qur'anic Education | 126 (31.7) |
| **Work in the Past 7 days** |  |
| Yes | 190 (47.5) |
| No | 210 (52.5) |

<sup>a</sup> Values are frequency (percentage) for categorical variables; median ± SD for continuous variables
<sup>b</sup> Values may not add to 100%

### Respondent Health Profile

Participant responses concerning perceptions of their personal health status, pre-existing health conditions, use of nicotine substances, and COVID-19 symptoms are displayed in **Table 2**. While 58% (n = 224) reported being in “good” health and only 3% (n = 13) reported using cigarettes or tobacco products, 50% (n = 197) (not shown in tables) of participants reported having one or more of the listed pre-existing conditions. In regards to symptoms being experienced, headache (49%, n = 188) and fever (23%, n = 88) were reported as the most common. Furthermore, 63% (n = 243) of respondents were concerned about getting adequate physical exercise, and 51% (n = 198) had concerns about job security (**Table 3**), indicating physical and financial precarities. Five percent (n = 21) of respondents also knew someone experiencing violence in their household (not shown in tables), and 27% (n = 106) anticipated contracting the virus at some point (not shown in tables). Additionally, 235 participants (59%) indicated that the nation-wide lockdown as a result of COVID-19 had decreased their income or caused them to lose their jobs when asked how COVID-19 had changed their daily lives (not shown in tables).

**Table 2.**
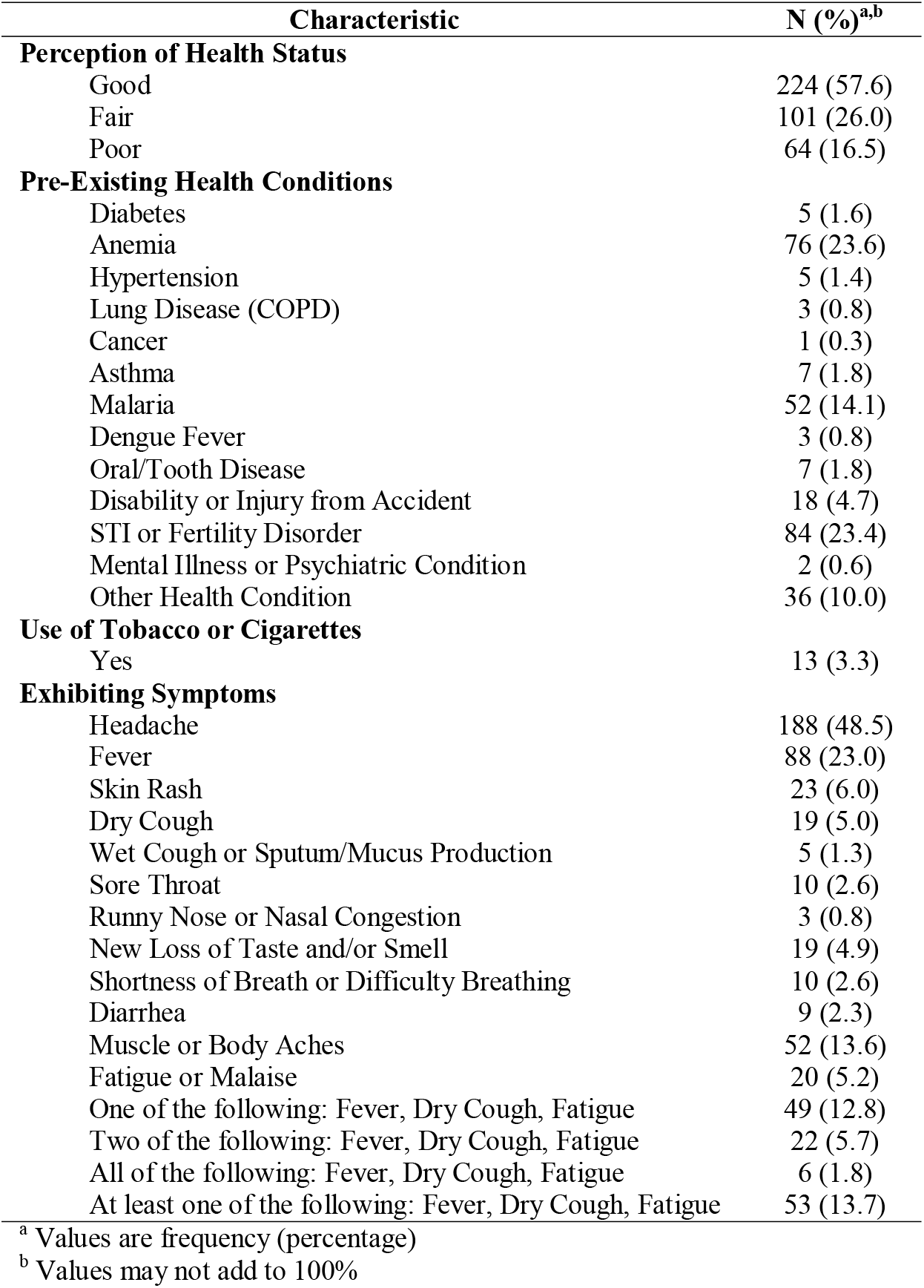
Respondent Health Profile.

| **Characteristic** | **N (%)<sup>a,b</sup>** |
| --- | --- |
| **Perception of Health Status** |  |
| Good | 224 (57.6) |
| Fair | 101 (26.0) |
| Poor | 64 (16.5) |
| **Pre-Existing Health Conditions** |  |
| Diabetes | 5 (1.6) |
| Anemia | 76 (23.6) |
| Hypertension | 5 (1.4) |
| Lung Disease (COPD) | 3 (0.8) |
| Cancer | 1 (0.3) |
| Asthma | 7 (1.8) |
| Malaria | 52 (14.1) |
| Dengue Fever | 3 (0.8) |
| Oral/Tooth Disease | 7 (1.8) |
| Disability or Injury from Accident | 18 (4.7) |
| STI or Fertility Disorder | 84 (23.4) |
| Mental Illness or Psychiatric Condition | 2 (0.6) |
| Other Health Condition | 36 (10.0) |
| **Use of Tobacco or Cigarettes** |  |
| Yes | 13 (3.3) |
| **Exhibiting Symptoms** |  |
| Headache | 188 (48.5) |
| Fever | 88 (23.0) |
| Skin Rash | 23 (6.0) |
| Dry Cough | 19 (5.0) |
| Wet Cough or Sputum/Mucus Production | 5 (1.3) |
| Sore Throat | 10 (2.6) |
| Runny Nose or Nasal Congestion | 3 (0.8) |
| New Loss of Taste and/or Smell | 19 (4.9) |
| Shortness of Breath or Difficulty Breathing | 10 (2.6) |
| Diarrhea | 9 (2.3) |
| Muscle or Body Aches | 52 (13.6) |
| Fatigue or Malaise | 20 (5.2) |
| One of the following: Fever, Dry Cough, Fatigue | 49 (12.8) |
| Two of the following: Fever, Dry Cough, Fatigue | 22 (5.7) |
| All of the following: Fever, Dry Cough, Fatigue | 6 (1.8) |
| At least one of the following: Fever, Dry Cough, Fatigue | 53 (13.7) |

<sup>a</sup> Values are frequency (percentage)<sup>b</sup> Values may not add to 100%

**Table 3.** Concerns Brought on by COVID-19.

| **Question** | **Yes N (%)<sup>a,b</sup>** | **No N (%)<sup>a,b</sup>** | **Not Sure N (%)<sup>a,b</sup>** |
| --- | --- | --- | --- |
| Getting adequate exercise/movement | 243 (62.6) | 135 (34.8) | 10 (2.6) |
| Mental health and wellbeing | 85 (22.1) | 244 (63.5) | 55 (14.3) |
| Job security | 198 (51.2) | 155 (40.1) | 34 (8.8) |
| Social distancing | 71 (18.7) | 292 (77.0) | 16 (4.2) |
| Social isolation in the case of infection | 53 (13.6) | 321 (82.5) | 15 (3.9) |
| Covering living expenses | 39 (10.0) | 277 (71.0) | 74 (19.0) |
| Caring for children | 36 (9.5) | 332 (87.6) | 11 (2.9) |
| The education of your child(ren) | 45 (11.6) | 330 (85.3) | 12 (3.1) |
| Social dynamics/conflict within the home | 37 (9.6) | 271 (70.0) | 79 (20.4) |
| Worried about getting COVID-19/Coronavirus | 259 (65.6) | 99 (25.1) | 37 (9.4) |
| Worried about loved ones getting COVID-19/Coronavirus | 265 (68.0) | 93 (23.9) | 32 (8.2) |

<sup>a</sup> Values are frequency (percentage)<sup>b</sup> Values may not add to 100%

### Knowledge of COVID-19

**Table 4** presents participants’ responses to true or false questions regarding COVID-19 susceptibility and preventative measures. Roughly 50% of responses to the question series were “I don’t know,” indicating a lack of information surrounding COVID-19 among IDPs. Most notably, the majority of participants displayed a lack of knowledge regarding the treatment of COVID-19, with about 79% of participants responding “I don’t know” when asked whether antibiotics could be used to treat or prevent COVID-19 (**Table 4**).

**Table 4.** Knowledge of COVID-19 disease and prevention.

| **Statement** | **Correct**<br>**N (%)<sup>a,b</sup>** | **Incorrect N**<br>**(%)<sup>a,b</sup>** | **Not Sure**<br>**N (%)<sup>a,b</sup>** |
| --- | --- | --- | --- |
| People showing no symptoms of being sick can spread a virus or contagious disease (True) | 150 (37.5) | 30 (7.5) | 220 (55.0) |
| It is possible to contract a virus or contagious disease by touching a surface or object that has the virus on it (True) | 191 (48.0) | 6 (1.5) | 201 (50.5) |
| Antibiotics can be used to treat COVID-19/Coronavirus (False) | 36 (9.1) | 51 (12.8) | 311 (78.1) |
| Antibiotics can be used to prevent infection from COVID-19 (False) | 37 (9.4) | 44 (11.2) | 313 (79.4) |
| People of all ages can become infected with COVID-19/Coronavirus (True) | 206 (51.8) | 19 (4.8) | 173 (43.3) |
| People of all racial, religious, and ethnic groups can become infected with COVID-19/Coronavirus (True) | 199 (50.4) | 17 (4.3) | 179 (45.3) |
| Eating garlic, ginger, black pepper, or lemon can lower your chances of getting infected with COVID-19/Coronavirus (False) | 12 (3.0) | 248 (62.8) | 135 (34.2) |
| Coronavirus spread from humans to humans, mainly through respiratory droplets (True) | 199 (50.6) | 13 (3.3) | 181 (46.1) |
| Coronavirus can also spread from feco-oral route (True) | 111 (28.1) | 34 (8.6) | 250 (63.3) |
| COVID-19/Coronavirus is just like the flu (False) | 20 (5.1) | 214 (54.0) | 162 (40.9) |
| Maintaining good personal hygiene and being socially responsible would help limit the spread of COVID-19/Coronavirus (True) | 252 (64.5) | 56 (14.3) | 83 (21.2) |
| Washing hands frequently using soap or the use of sanitizer would help limit the spread of COVID-19/Coronavirus (True) | 253 (64.9) | 55 (14.1) | 82 (21.0) |
| Avoiding handshaking behavior would help limit the spread of COVID-19/Coronavirus (True) | 219 (56.7) | 65 (16.8) | 102 (26.4) |
| Avoiding placing fingers into eyes, nose, and mouth would help limit the spread of COVID-19/Coronavirus (True) | 181 (47.0) | 74 (19.2) | 130 (33.8) |
| Coughing and sneezing into the elbow or within the clothing is a good practice in helping to limit the spread of COVID-19/Coronavirus (True) | 209 (54.4) | 64 (16.7) | 111 (28.9) |
| Limiting eating meat, eggs, and fishes would help limit the spread of COVID-19/Coronavirus (False) | 153 (39.8) | 116 (30.2) | 115 (30.0) |
| Following social distancing measures and avoiding crowded places would help limit the spread of COVID-19/Coronavirus (True) | 172 (44.7) | 76 (19.7) | 137 (35.6) |
| For someone without any symptoms of COVID-19/Coronavirus, wearing a face mask is considered an appropriate and protective measure against COVID-19/Coronavirus (True) | 144 (37.4) | 100 (26.0) | 141 (36.6) |
| Proper usage of face mask during an outbreak should include covering nose, mouth, and chin with the colored side facing outside (True) | 167 (43.4) | 64 (16.6) | 154 (40.0) |
| Staying at home would play a significant role in helping to limit the spread of COVID-19/Coronavirus (True) | 215 (56.0) | 84 (21.9) | 85 (22.1) |
| A person with pre-existing medical conditions such as heart diseases, diabetes, hypertension, and cancer are at greater risk of COVID-19/Coronavirus related infection (True) | 172 (44.7) | 64 (16.6) | 149 (38.7) |
| COVID-19/Coronavirus can only infect old people (False) | 195 (50.9) | 69 (18.0) | 119 (31.1) |
| There is no chance of survival once the person is infected with COVID-19/Coronavirus (False) | 195 (52.3) | 67 (18.0) | 111 (29.8) |

<sup>a</sup> Values are frequency (percentage)<sup>b</sup> Values may not add to 100%

Furthermore, the majority of participants marked that they did not know the current possible treatment (83%, n = 327), the symptoms (50%, n = 201), or the incubation period of COVID-19 (79%, n = 313) (all not shown in tables). Another 60% (n = 237) reported being unfamiliar with the practice of social distancing (not shown in tables), and 63% (n = 250) did not know that asymptomatic individuals could spread the virus (**Table 4**). However, a sizable proportion of participants were able to recognize headache (34%, n = 135), fever (45%, n = 179), cough (35%, n = 139), and shortness of breath or difficulty breathing (31%, n = 12) as symptoms of COVID-19 (not shown in tables). Still, the proportion of participants who were not familiar with basic background information surrounding COVID-19 was higher than the proportion who answered questions correctly.

When respondents were asked what role religion played in the COVID-19 outbreak, approximately 34% wrote in short answer responses that religion played an “important” or “very important” role and another 22% specified that reading the Qur’an was of particular importance (not shown in tables). Additionally, 49% of the participants (n = 190) reported that they trusted religious officials to provide information about COVID-19, while only 37% (n = 144) and 5% (n = 19) said that they similarly trusted health officials and humanitarian aid workers, respectively (**Table 5**). In contrast, 90% (n = 360) reported accessing traditional media services (radio, television, and newspapers), with only radio being indicated as a more trustworthy source than religious officials (57%, n = 225). Additionally, only 4% (n = 11) of respondents stated that they would go to health information providers in IDP camps if they suspected they had contracted the virus (not shown in tables). As a majority of respondents indicated that they felt that healthcare workers and the government were working together “moderately” or “a lot” to prevent the spread of COVID-19 (**Table 6**), participants’ lack of initiative to approach health care workers seems to indicate a lack of COVID-19 related information and services in refugee camps instead of a distrust of such.

**Table 5.** Respondent Trusted Sources of Information Surrounding COVID-19.

| Source | N (%)<sup>a</sup> |
| --- | --- |
| **Sources of Information** |  |
| News, Media (e.g. TV, Radio, Newspapers) | 360 (90.0) |
| Informational calls/SMS | 161 (40.3) |
| Social Media (e.g. Facebook, Twitter, WhatsApp, YouTube, Instagram, Snapchat) | 21 (5.3) |
| Official Government/International Websites (e.g. MoH, DoH, WHO, CDC) | 10 (2.5) |
| Family Members, Colleagues, Friends | 38 (10.0) |
| Employer, Work Colleagues, and Others at Work | 7 (1.8) |
| Non-Governmental Organizations (NGOs) | 21 (5.3) |
| Informational Campaigns | 17 (4.3) |
| Local or Community Leaders | 8 (2.0) |
| Journals | 8 (2.0) |
| **Trust in Sources to Provide COVID-19 Information** |  |
| Family Members | 162 (41.3) |
| Religious Official | 190 (48.5) |
| Community Elder | 31 (8.0) |
| Health Official | 144 (36.7) |
| Humanitarian Aid Worker | 19 (4.8) |
| Social Media | 17 (4.3) |
| Web News | 9 (2.3) |
| Television | 41 (10.5) |
| Radio | 225 (57.4) |

<sup>a</sup> Values are frequency (percentage)

**Table 6.**
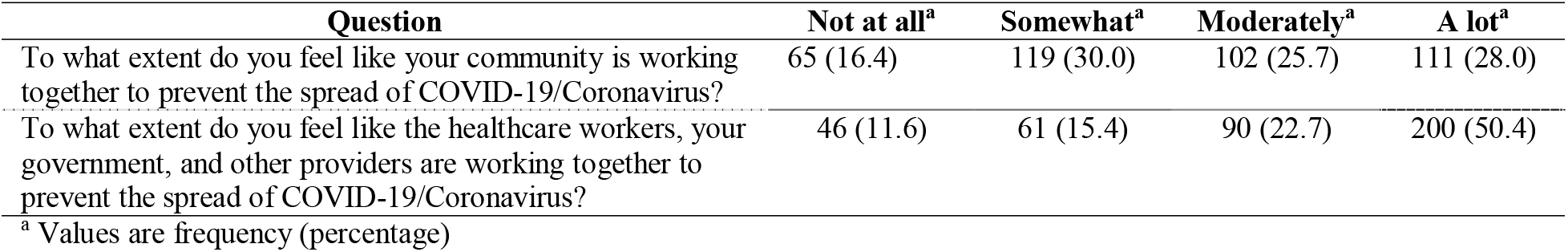
Faith in Community Action Against COVID-19.

### Access to Treatment and Preventative Services

**Table 7** reports participants’ access to essential services, preventative resources, and treatments for COVID-19. Thirty-four percent (n = 135) of respondents reported not being able to buy essential food items in the last week and, of those, 74% (n = 100) reported having either to skip and eat smaller meals or buy lower quality food (not shown in tables). A majority lacked ease of access to washing facilities (71%, n = 277), soap (67%, n = 256), disinfectants (98%, n = 382), and face masks (89%, n = 373). Although respondents lacked access to such resources, they still reported carrying out basic preventative procedures such as washing hands more often (74%, n = 292) and avoiding shaking hands (57%, n = 222) (not shown in tables). While only 5% (n = 20) and 25% (n = 98) of participants reported using more disinfectants and practicing social distancing, 77% (n = 296) took at least one preventative action (not shown in tables).

Additionally, 185 (47%) respondents indicated that camp living conditions needed to change to prevent the spread of COVID-19 (not shown in tables). In short answer responses, 142 (80%) respondents expressed desires for improved sanitation, hygiene, or housing conditions, and 14 (8%) specifically indicated their desire for more stringent social distancing practices (not shown in tables). Maintaining physical space from others was also reported to be difficult, as 40% (n = 159) and 32% (n = 128) indicated that it be impossible or only somewhat possible to self-isolate if taken ill, respectively (not shown in tables). An additional 10 (6%) respondents indicated their desire for the distribution of more information regarding COVID-19 (not shown in tables).

Similarly, respondents’ self-reported access to COVID-19 screening and medical services was also low, with 97% (n = 381) saying that they could not access screening and only 20% (n = 79) stating that they were “Strongly Confident” or “Moderately Confident” that they could receive medical services if infected (**Table 7**). However, most participants had received a vaccination for any condition within the past few years (59%, n = 232) (not shown in tables), and a further 54% (n = 215) responded “Moderately” or “A lot” when asked how available vaccinations were to them, suggesting that COVID-19 specific services would be harder to access. Participants also reported accessibility issues, with 48% (n = 191) stating that there was no healthcare facility nearby and, of those, 31% (n = 61) said that it would take over an hour to reach it (not shown in tables).

**Table 7.** Essential Resources for COVID-19 Response.

| **Question** | **Yes N (%)<sup>a,b</sup>** | **No N (%)<sup>a,b</sup>** |
| --- | --- | --- |
| Access to Clean Water or a Clean Water Source | 362 (91.6) | 33 (8.4) |
| Access to Washing Facilities | 113 (29.0) | 277 (71.0) |
| Access to Soap | 129 (33.6) | 256 (66.5) |
| Access to Disinfectants, such as Hand Sanitizers and Cloth Wipes | 8 (2.1) | 382 (98.0) |
| Ability to Buy Essential Food Items | 261 (65.9) | 135 (34.1) |
| Access to Face Masks | 43 (10.9) | 351 (89.1) |
| Ability to Maintain Clean and Sanitary Conditions at Home | 325 (81.9) | 69 (17.4) |
| Access to Nearby Healthcare Facility | 194 (49.0) | 191 (48.2) |
| Confident in Ability to Access Healthcare Facility if Infected with COVID-19 | 79 (20.0) | 275 (69.4) |
| Access to Vaccines | 215 (54.2) | 174 (43.8) |
| Access to COVID-19/Coronavirus Screening | 6 (1.5) | 381 (96.9) |

<sup>a</sup> Values are frequency (percentage)<sup>b</sup> Values may not add to 100%

### Bi- and Multivariate Analyses Examining Concern about COVID-19 Contraction

Lastly, in an adjusted model (**Table 8**), those who were concerned about contracting COVID-19 were more likely to perceive their current health status as fair, describe an increased presence of anxiety or stress, and introduce behavioral changes to protect themselves against the virus compared to those who weren’t concerned with contracting COVID-19. Compared to those who perceived their current health status as good, those who perceived their health status to be fair showed 5.69 (95% CI: 2.36–13.71) times higher odds of being concerned about contracting COVID-19, while those who felt more anxious or nervous and those who introduced one behavioral change showed 10.16 (95% CI: 4.83–21.36) and 5.20 (95% CI: 1.99–13.58) times increased odds of being concerned about disease contraction, respectively. In addition, a dose-response relationship in the number of distancing and sanitization behavioral changes implemented was observed, with those who introduced two or more behavioral changes showing 9.89 (95% CI: 4.88–20.02) times higher odds of being concerned about contracting COVID-19 when compared with those without any introduced behavioral changes. Lastly, the older half of our sample (older than 32 years of age) showed 1.93 (95% CI: 1.06 – 3.51) times higher odds in being concerned about contracting COVID-19.

**Table 8.** Bivariate and multivariable associations between study variables and concern regarding COVID-19 contraction.

| Characteristic | N<sup>a</sup><br>(40<br>1) | N(%)<br>concerned<br>about<br>COVID<br>contraction | Unadjusted OR (95%<br>CI) | Un-<br>adjusted<br>p-value | Adjusted OR<br>(95% CI)<sup>b</sup> | Adjusted<br>p-value |
| --- | --- | --- | --- | --- | --- | --- |
| **Gender** |  |  |  |  |  |  |
| Male | 52 | 38 (73.1) | 1.00 |  | 1.00 |  |
| Female | 304 | 221 (72.7) | 0.98 (0.51 – 1.90) | 0.955 | - | - |
| **Age (years)** |  |  |  |  |  |  |
| 18-32 | 182 | 122 (67.0) | 1.00 |  | 1.00 |  |
| 33-80 |  |  | 1.73 (1.08 – 2.77) | 0.023 | 1.93<br>(1.06 – 3.51) | 0.032 |
|  | 176 | 137 (77.8) |  |  |  |  |
| **Formal Education** |  |  |  |  |  |  |
| 0 years | 314 | 229 (72.9) | 1.00 |  | 1.00 |  |
| 1+ years | 43 | 29 (67.4) | 0.77 (0.39 – 1.53) | 0.452 | - | - |
| **Qur'anic Education** |  |  |  |  |  |  |
| No | 245 | 177 (72.2) | 1.00 |  | 1.00 |  |
| Yes | 112 | 81 (72.3) | 1.00 (0.61 – 1.65) | 0.988 | - | - |
| **Currently Employed** |  |  |  |  |  |  |
| No | 184 | 127 (69.0) | 1.00 |  | 1.00 |  |
| Yes | 174 | 132 (75.9) | 1.41 (0.88 – 2.25) | 0.149 | - | - |
| **Current Health Perception** |  |  |  |  |  |  |
| Good | 202 | 137 (67.8) | 1.00 |  | 1.00 |  |
| Fair |  |  | 4.57 (2.19 – 9.53) | <0.001 | 5.69<br>(2.36 – 13.71) | <0.001 |
|  | 88 | 79 (89.8) |  |  |  |  |
| Poor |  |  | 0.49 (0.27 – 0.87) | 0.016 | 0.57<br>(0.25 – 1.31) | 0.185 |
|  | 58 | 34 (58.6) |  |  |  |  |
| **Pre-existing Conditions** |  |  |  |  |  |  |
| 0 | 175 | 128 (73.1) | 1.00 |  | 1.00 |  |
| 1 |  |  | 1.27 (0.76 – 2.11) | 0.370 | 0.80<br>(0.39 – 1.62) | 0.531 |
|  | 110 | 83 (75.5) |  |  |  |  |
| 2+ |  |  | 0.66 (0.38 – 1.15) | 0.140 | 0.50<br>(0.23 – 1.10) | 0.084 |
|  | 72 | 47 (65.3) |  |  |  |  |
| **Tobacco Use** |  |  |  |  |  |  |
| No | 340 | 245 (72.1) | 1.00 |  | 1.00 |  |
| Yes | 11 | 8 (72.7) | 1.03 (0.27 – 3.98) | 0.962 | - | - |
| **Present COVID-19 symptoms** |  |  |  |  |  |  |
| 0 | 153 | 110 (71.9) | 1.00 |  | 1.00 |  |
| 1 |  |  | 0.60 (0.36 – 1.01) | 0.053 | 0.60<br>(0.30 – 1.21) | 0.153 |
|  | 91 | 59 (64.8) |  |  |  |  |
| 2+ |  |  | 1.80 (1.05 – 3.08) | 0.034 | 1.73<br>(0.83 – 3.64) | 0.147 |
|  | 111 | 89 (80.2) |  |  |  |  |
| **Received COVID Info** |  |  |  |  |  |  |
| None / Somewhat | 262 | 186 (71.0) | 1.00 |  | 1.00 |  |
| Moderately / A lot | 96 | 73 (76.0) | 1.30 (0.76 – 2.22) | 0.345 | - | - |
| **New Stress or Anxiety** |  |  |  |  |  |  |
| No | 54 | 21 (38.9) | 1.00 |  | 1.00 |  |
| Yes |  |  | 5.68 (3.08 – 10.48) | <0.001 | 10.16<br>(4.83 – 21.36) | <0.001 |
|  | 300 | 235 (78.3) |  |  |  |  |
| **New actions to protect self** |  |  |  |  |  |  |
| 0 | 78 | 39 (50.0) | 1.00 |  | 1.00 |  |
| 1 |  |  | 1.25 (0.64 – 3.44) | 0.520 | 5.20 |  |
|  | 54 | 41 (75.9) |  |  | (1.99 – 13.58) | <0.001 |
| 2+ |  |  | 2.49 (1.54 – 4.01) | <0.001 | 9.89 |  |
|  | 222 | 176 (79.3) |  |  | (4.88 – 20.02) | <0.001 |
| **Belief that camp needs to be improved to handle COVID** |  |  |  |  |  |  |
| No | 171 | 115 (67.3) | 1.00 |  | 1.00 |  |
| Yes | 181 | 141 (77.9) | 1.72 (1.07 – 2.76) | 0.026 | - | - |
| **Past Vaccinations** |  |  |  |  |  |  |
| No | 131 | 95 (72.5) | 1.00 |  | 1.00 |  |
| Yes | 216 | 157 (72.7) | 1.01 (0.62 – 1.64) | 0.973 | - | - |

<sup>a</sup> Numbers may not sum to total due to missing data.
<sup>b</sup> For the fully-adjusted model, N = 341

## Discussion

This study is the first to explore knowledge of COVID-19, access to essential services, COVID- 19 symptoms and risk factors among individuals living in IDP camps in Somalia. To our knowledge, it is also one of two studies that document knowledge of COVID-19 and prevalence of COVID-19 symptoms and risk factors among displaced populations globally, with the other study conducted in Cox’s Bazar, Bangladesh[27]. Overcrowding, limited access to WASH services, and a lack of health infrastructure leave displaced populations living in IDP camps particularly vulnerable to the spread of infectious disease such as COVID-19[2].

### Respondent Health Profile

Although a majority of our sample (57.6%) reported that they were in good health, a considerable proportion of our sample reported experiencing symptoms that could potentially be attributed to COVID-19, including headache (48.5%), fever (23.0%), or muscle or body aches (13.6%)[29]. Previous studies on health profiles of displaced populations or individuals living in such settings are unavailable in Somalia and are sparse in international settings, limiting the potential for cross-comparison. However, compared to a similar study conducted in refugee camps in Bangladesh, our sample exhibited higher proportions of individuals experiencing fevers, headaches, shortness of breath, loss of taste or smell, and muscle or body aches[27]. In addition, our study findings reinforce concerns put forward in the literature regarding the spread of COVID-19 in humanitarian settings, with participants reporting concerns including the following: getting adequate exercise (62.6%), their mental health and well-being (22.1%), job security (51.2%), practicing social distancing (18.7%), experiencing conflict within the home (9.6%), and their ability to buy essential foods in the last 7 days (65.9%). These concerns could serve as priority areas for health and social service providers to engage with and address, particularly food insecurity, job security, mental health, and physical exercise.

Only 3.3% of our sample reported use of tobacco or cigarettes, which appears to be consistent with smoking prevalence in Somalia, especially considering that the majority of our sample consisted of women[30]. That being said, our study reports a lower rate of tobacco use when compared to other studies conducted in similar limited-resource settings[31]. Furthermore, though little information is available regarding the health conditions of individuals living in IDP camps, self-reported pre-existing conditions from our sample appear to be considerably low, particularly for chronic and mental health conditions, when compared to values expected from the literature[32–35]. Health staff administering surveys reported that many participants did not know what the surveyed conditions were, had little recollection of previous diagnoses, and had rarely visited a health professional in the past, which would be consistent with the significant lack of formal education (88.7%) among participants and lack of access to a nearby healthcare facility (48.2%) and may explain the lower reported prevalence of chronic, pre-existing conditions found in this study.

### Knowledge and Perceptions of COVID-19

This study found very poor knowledge related to COVID-19 and relevant universal safety measures among individuals living in IDP camps, with significant knowledge gaps in COVID-19 symptoms, treatment, and prevention measures such as social distancing. Though there is limited literature on this topic, especially in camp-like or humanitarian contexts, our sample exhibited significantly poorer knowledge of COVID-19 and safety measures, as compared to studies conducted in China, the United States, the United Kingdom, and Nepal[36, 37, 26]. And, although related literature in humanitarian settings is severely limited, this discrepancy persists among the sparse COVID-related published research in camp-like settings, such as that of a study conducted in Cox’s Bazar, Bangladesh[27]. These findings may also be attributed to the lack of formal education (88.7%) among study participants, as well as the reality that Somalia has some of the lowest health indicators in the world and low levels of health literacy stemming from decades of conflict, civil strife, and limited access to healthcare and education – particularly for displaced populations like IDPs[38–41].

The older half of our sample, those who had implemented at least one COVID-19-related behavior change, and those who characterize their health as “fair” were significantly more likely to be concerned about getting infected by COVID-19. While these results demonstrate possible increased apprehension by those who are physically or mentally vulnerable to the changes imposed by the COVID-19 pandemic, our data interestingly demonstrate that those who characterized their health as “fair” were significantly more likely to be concerned about COVID- 19 contraction, while those who characterize their health as “poor” were less likely, albeit insignificantly. These results may be explained by a small sample size, as only 58 respondents perceived their health status as “poor.”

The importance of education as a preventative measure in controlling the transmission of COVID-19 in camp settings cannot be understated. Given significant gaps in COVID-19 knowledge among Somali IDP camp residents, instituting precautionary measures, such as public awareness, is paramount. Knowledge is a critical determinant of health behaviors, and through targeted educational interventions, IDP camp residents can play an essential role in preventing community spread of the virus among those displaced and not displaced[42]. Ninety percent of our study sample utilized news and media outlets, such as the radio, to access information about COVID-19, and 40.3% used informational calls or text messages. Using such platforms to educate camp residents may be effective in disseminating information about COVID-19 prevention and treatment. Similarly, trusted sources, such as religious officials (48.5%), health officials (36.7%), or the radio (57.4), should be mobilized to engage with community members and to dispel misinformation about COVID-19.

### Access to Treatment and Preventative Services

This study’s results reveal a significant lack of access to treatment and preventative services for COVID-19 among individuals living in IDP camps, which is consistent with other assessments of access to health and water, hygiene, and sanitation services in Somalia and in camp-like settings in other countries[2, 43–45]. Of particular note, only 1.5% of participants reported having access to COVID-19 screening tests, and 43.8% reported not having access to any vaccines which may inhibit adequate uptake of a future COVID-19 vaccine. In addition, these findings reflect not only the lack of access to services necessary to prevent COVID-19 transmission and to identify and treat existing cases, but also the lack of access to services that may exacerbate known comorbidities of COVID-19, such as hypertension, cancer, cardiovascular disease, and diabetes, which are increasingly difficult to provide continuous care for during the COVID-19 pandemic[1, 4]. Generally, respondents reported an inability to participate in preventative measures due to a lack of material resources such as face masks and disinfectants, lack of information about COVID-19 and such measures, and inadequate living conditions to enforce social distancing protocols. Additionally, respondents reported limited confidence that they would be able to be treated if they contracted the virus. These findings reinforce the necessity for substantial investment in improving access to water, sanitation, and hygiene services, as well as healthcare services, for displaced populations living in camps.

## Limitations

This study’s sampling methodology is subject to several limitations. First, participants were recruited from twelve Somali IDP camps across six areas (Ceelasha, Lafoole, Xaawo Cabdi, Carbiska, and Afgooye) of the Lower Shabelle region, which limits the generalizability of the study findings to IDP camps in other areas of Somalia. That being said, the Lower Shabelle region harbors one of the largest IDP populations in Somalia, as well as one of the highest concentrations of individuals in need of humanitarian support[46]. Second, because an overwhelming majority of participants surveyed (86.2%) were women, the generalizability of the study results to males living in IDP camps may be limited. Third, the respondent health profiles reported (**Table 2**) consist exclusively of self-reported data. As such, the limited reliability of participant responses for pre-existing conditions must be noted. Fourth, because our multivariate analysis did not adjust for potential confounding factors, residual confounding may have been introduced. Finally, given that this study utilizes a cross-sectional survey design, it cannot be used to analyze the evolution of COVID-19 knowledge, health conditions, and access to services in Somali IDP camps over an extended period of time.

## Conclusion

Ultimately, this study provides strong evidence for immense gaps in knowledge and perceptions of COVID-19 and access to treatment and preventative services among individuals living in Somali IDP camps. In addition, this study also provides insight into the health profile of IDP camp residents, as well as their concerns during the COVID-19 pandemic. A massive influx of additional resources and targeted interventions will be required to adequately address COVID-19 in Somalia, starting with educating those individuals most vulnerable to infection. Future research is needed to further elucidate the health profile of camp residents and the most at-risk individuals and to identify entry points to facilitate the implementation of COVID-19 preventative and treatment measures. International organizations operating within Somalia, as well as the Somali government, should view investments in addressing COVID-19 as long-term investments in sustainable health infrastructure, as well as the health and prosperity of its population.

## Data Availability

All data generated or analyzed during this study are included in this article.

## Declarations

Ethics approval and consent to participate: This study received approval from the ethics board at SIMAD University in Somalia and was deemed exempt from review by the Yale IRB (ID #2000028344). Verbal consent to participate in the study was obtained privately from each participant prior to the survey being interviewer-administered by trained staff from the Hagarla Institute. If a participant consented, they were informed of their right to withdraw at any time without any consequences and were presented with a brief overview of the study before any survey questions were administered. All information collected was confidential and anonymized.

### Consent for publication: Not applicable

Patient and Public Involvement: The research questions identified and explored in this study were informed by concerns raised by Somali community members arising from COVID-19. In consultation with community members, as well as community organizers, the research team designed the survey tool and highlighted key research priorities. Relevant results from this study will be disseminated to community members, operating organizations within the area, and governmental agencies to facilitate and improve current actions to address COVID-19 among those displaced within Somalia.

### Availability of data and materials

All data generated or analyzed during this study are included in this published article.

### Competing interests

The authors declare that they have no competing interests. Funding: Not applicable

### Authors’ contributions

JA, SA, LW, KK, AA, and DM contributed to the design of the study. AA and DM contributed to the collection of data. JA, SA, LW, EW, and NH each contributed to the analysis of data. All authors contributed to the interpretation of data and the development of the manuscript.

## Acknowledgements

We would like to thank the Hagarla Institute and the following individuals for their support of our research: Mohamud Abdi Mohamed, Mohamud Abdinasir Farah, Mahad Mahamed Jamac, Mahad Abdirishid Warsame, Aini Mohamed Abdi, Zainab Hussein Ibrahim, Faiza Ibrahim Mohamed, Shukri Mohamud Mahdi, Ahmed Abdukhadir Shacir, Mukhtar Mohamed Mohamud, Ali Mukhtar Ahmed, Mohamed Ahmed Hassan, Zeinab Mahmud Derow.

## Notes

### Competing Interest Statement

The authors have declared no competing interest.

### Funding Statement

No external funding was used for this project.

### Author Declarations

This study received approval from the ethics board at SIMAD University in Somalia and was deemed exempt from review by the Yale IRB (ID #2000028344).

## References

1 Hopman J, Allegranzi B, Mehtar S. Managing COVID-19 in Low-and Middle-Income Countries. JAMA 2020;323(16):1549–1550. doi:10.1001/jama.2020.4169

2 Kluge HHP, Jakab Z, Bartovic J, et al. Refugee and migrant health in the COVID-19 response. The Lancet 2020;395(10232):1237–9.

3 Global Trends: Forced Displacement in 2017 [Internet]. UNHCR The UN Refugee Agency. UNHCR. 2018 [cited 2020Jul13]. Available from: https://www.unhcr.org/5b27be547.pdf

4 Lau LS, Samari G, Moresky RT, et al. COVID-19 in humanitarian settings and lessons learned from past epidemics. Nature Medicine 2020Apr8;26(5):647–8.

5 Home [Internet]. MOH. 2020 [cited 2020Jul8]. Available from: https://moh.nomadilab.org/

6 Somalia faces another desperate situation as COVID-19 cases spiral, IRC calls for increased support to save lives [Internet]. International Rescue Committee (IRC). 2020 [cited 2020Jul8]. Available from: https://www.rescue.org/press-release/somalia-faces-another-desperate-situation-covid-19-cases-spiral-irc-calls-increased

7 Somalia | COVID-19 Response. UNHCR. 28 April 2020 [cited 2020Jul8]. Available from: https://reporting.unhcr.org/sites/default/files/UNHCR%20Somalia%20COVID-19%20Response%20Update%20-%2028APR20_0.pdf

8 Ali A, Handuleh J, Patel P, et al. The most fragile state: healthcare in Somalia. Medicine, Conflict and Survival2014;30(1):28–36. doi:10.1080/13623699.2014.874085

9 Warsame A, Handuleh J, Patel P. Prioritization in Somali health system strengthening: a qualitative study. Int Health 2016;8(3):204–210. doi:10.1093/inthealth/ihv060

10 Somalia Responds: Together we can fight COVID-19 in Somalia [Internet]. Somalia Responds | International Organization for Migration. [cited 2020Jul8]. Available from: https://www.iom.int/donate/campaigns/somalia-responds

11 World Health Organization Humanitarian Response Plans in 2015: Somalia [Internet]. World Health Organization. 2015 [cited 2020Jul13]. Available from: https://www.who.int/hac/donorinfo/somalia.pdf

12 Elkheir N, Sharma A, Cherian M, et al. A cross-sectional survey of essential surgical capacity in Somalia. BMJ Open 2014;4:e004360. doi:10.1136/bmjopen-2013-004360

13 Gele AA, Ahmed MY, Kour P, et al. Beneficiaries of conflict: a qualitative study of people’s trust in the private health care system in Mogadishu, Somalia. Risk Management and Healthcare Policy 2017;10:127–135. doi:10.2147/rmhp.s136170

14 Somalia [Internet]. World Health Organization. 2020 [cited 2020 Jul 8]. Available from: https://www.who.int/countries/som/en/

15 Soliman AA, Abdulrahman BMA, Ali JA, et al. Water associated diseases amongst children in IDPs camps and their relation to family economics status: case study of Abuschock IDPs camp, North Darfur State, Sudan. International Journal of Research – Granthaalayah2017;5(4):214–227. https://doi.org/10.5281/zenodo.571528

16 Global Health Security Index: Building Collective Action and Accountability p. 274, “Somalia.” [Internet]. 2019 [cited 2020Jul8]. Available from: https://www.ghsindex.org/wp-content/uploads/2020/04/2019-Global-Health-Security-Index.pdf

17 Operational Portal. Document – CCCM COVID-19 Contingency Plan [Internet]. UNHCR. 2020 [cited 2020Jul8]. Available from: https://data2.unhcr.org/en/documents/details/75050

18 Kumamaru K, Khayre O, Ito C. Improving access to safe water for internally displaced persons (IDPs) in a fragile state, Somalia [Internet]. Loughborough University; 2013 [cited 2020Jul13]. Available from: https://hdl.handle.net/2134/30919

19 Somalia: Critical juncture to curb spread of COVID-19 and save lives [Internet]. International Committee of the Red Cross. 2020 [cited 2020Jul8]. Available from: https://www.icrc.org/en/document/somalia-critical-juncture-curb-spread-covid-19-and-save-lives

20 Truelove SA, Abrahim O, Altare C, et al. The Potential Impact of COVID-19 in Refugee Camps in Bangladesh and Beyond: a modeling study. PLOS Medicine 2020. doi:10.1101/2020.03.27.20045500

21 Bigg MM. Support for internally displaced people needs to be ‘urgently stepped up’ [Internet]. UNHCR 2020 [cited 2020Jul8]. Available from: https://www.unhcr.org/en-us/news/latest/2020/4/5e9839e74/support-internally-displaced-people-needs-urgently-stepped.html

22 COVID-19: Do not forget internally displaced persons, UN expert urges Governments worldwide [Internet]. United Nations Human Rights Office of the High Commissioner. OHCHR; 2020 [cited 2020Jul8]. Available from: https://www.ohchr.org/EN/NewsEvents/Pages/DisplayNews.aspx?NewsID=25763

23 Coronavirus [Internet]. World Health Organization. World Health Organization; 2020 [cited 2020Jul8]. Available from: http://www.who.int/health-topics/coronavirus

24 Symptoms of Coronavirus [Internet]. Centers for Disease Control and Prevention. Centers for Disease Control and Prevention; 2020 [cited 2020Jul13]. Available from: http://www.cdc.gov/coronavirus/2019-ncov/symptoms-testing/symptoms.html

25 Wesolek P, Sydney C. Edited by Martha Crowley. Assessing the severity of displacement. Internal Displacement Monitoring Centre. 2020 [cited 2020Jul13]. Available from: http://www.internal-displacement.org/sites/default/files/publications/documents/Severity%20Report%202019.pdf

26 Singh, D.R., Sunuwar, D.R., Karki, K. et al. Knowledge and Perception Towards Universal Safety Precautions During Early Phase of the COVID-19 Outbreak in Nepal. J Community Health2020. https://doi.org/10.1007/s10900-020-00839-3

27 Lopez-Pena P, Austin Davis C, Mushfiq Mobarak A, et al. Prevalence of COVID-19 symptoms, risk factors, and health behaviors in host and refugee communities in Cox’s Bazar: a representative panel study. [Preprint]. Bull World Health Organ. E-pub: 11 May 2020. doi:http://dx.doi.org/10.2471/BLT.20.265173

28 Qualtrics L. Qualtrics [software]. Utah, USA: Qualtrics.2014.

29 Huang C, Wang Y, Li X, et al. Clinical features of patients infected with 2019 novel coronavirus in Wuhan, China. The Lancet 2020;395(10223):497–506.

30 GBD 2015 Tobacco Collaborators. Smoking prevalence and attributable disease burden in 195 countries and territories, 1990–2015: a systematic analysis from the Global Burden of Disease Study 2015. The Lancet2017;389(10082):1885–906.

31 Jawad M, Khader A, Millett C. Differences in tobacco smoking prevalence and frequency between adolescent Palestine refugee and non-refugee populations in Jordan, Lebanon, Syria, and the West Bank: cross-sectional analysis of the Global Youth Tobacco Survey. Conflict and Health2016;10(1).

32 Jong JPD, Scholte WF, Koeter MWJ, et al. The prevalence of mental health problems in Rwandan and Burundese refugee camps. Acta Psychiatrica Scandinavica 2000;102(3):171–7.

33 Doocy S, Lyles, E, Roberton T, et al. Prevalence and care-seeking for chronic diseases among Syrian refugees in Jordan. BMC Public Health 2015;15(1097). https://doi.org/10.1186/s12889-015-2429-3

34 Amara A, Aljunid S. Noncommunicable diseases among urban refugees and asylum-seekers in developing countries: a neglected health care need. Global Health 2014;10(1):24. https://doi.org/10.1186/1744-8603-10-24

35 Alawa J, Maiky C, Khoshnood K, Fouad FM. Cancer prevention and treatment in humanitarian settings: an urgent and unmet need. The Lancet Oncology 2019;20(12):1635–6.

36 Zhong B-L, Luo W, Li H-M, et al. Knowledge, attitudes, and practices towards COVID-19 among Chinese residents during the rapid rise period of the COVID-19 outbreak: a quick online cross-sectional survey. International Journal of Biological Sciences 2020;16(10):1745–52. https://doi.org/10.7150/ijbs.45221

37 Geldsetzer P. Knowledge and perceptions of COVID-19 among the general public in the United States and the United Kingdom: A Cross-sectional Online Survey. Annals of Internal Medicine 2020. https://doi.org/10.7326/M20-0912

38 Klutse V. Measuring Health Literacy Among Somali Men Over the Age of 45 – A Pilot Study. Master of Science in Nursing Theses 2014;12. http://digitalcommons.cedarville.edu/nursing_theses/12

39 Refugee Health Profiles [Internet]. Centers for Disease Control and Prevention. Centers for Disease Control and Prevention; 2018 [cited 0Jul11]. Available from: https://www.cdc.gov/immigrantrefugeehealth/profiles/somali/background.html

40 Ferris E, Winthrop R. Education and Displacement: Assessing Conditions for Refugees and Internally Displaced Persons affected by Conflict [Internet]. Humanitarian Library. The Brookings Institution; 2010 [cited 2020Jul8]. Available from: https://www.humanitarianlibrary.org/sites/default/files/2014/02/190715e.pdf

41 Drumtra J. Internal Displacement In Somalia [Internet]. The Brookings Institution. 2014 [cited 2020Jul8]. Available from: https://www.refworld.org/pdfid/54bd197b4.pdf

42 Kenkel DS. Health behavior, health knowledge, and schooling. Journal of Political Economy 1991;99(2):287–305. https://doi.org/10.1086/261751

43 Ivanova O, Rai M, Kemigisha E. A Systematic Review of Sexual and Reproductive Health Knowledge,Experiences and Access to Services among Refugee, Migrant and Displaced Girls and Young Women in Africa. International Journal of Environmental Research and Public Health2018;15(8).

44 Pandemic influenza preparedness and mitigation in refugee and displaced populations. WHO guidelines for humanitarian agencies [Internet]. World Health Organization. World Health Organization; 2008 [cited 2020July13]. Available from: https://www.who.int/csr/resources/publications/swineflu/pandemic_preparedness_refugee/en/

45 Blundell H, Milligan R, Norris SL, et al. WHO guidance for refugees in camps: systematic review. BMJ Open 2019;9(9):1–7. DOI: 10.1136/bmjopen-2018-027094.

46 2019 Humanitarian Needs Overview: Somalia [Internet]. ReliefWeb. UNOCHA; 2018 [cited 2020Jul12]. Available from: https://reliefweb.int/sites/reliefweb.int/files/resources/Somalia_2019_HNO.PDF

